# Sedentary lifestyle, Physical Activity, and Gastrointestinal Diseases: Evidence from Mendelian Randomization Analysis

**DOI:** 10.1101/2023.03.15.23287301

**Authors:** Jie Chen, Xixian Ruan, Tian Fu, Shiyuan Lu, Dipender Gill, Zixuan He, Stephen Burgess, Edward L Giovannucci, Susanna C. Larsson, Minzi Deng, Shuai Yuan, Xue Li

## Abstract

**Objectives:** The causal associations of physical activity and sedentary behavior with the risk of gastrointestinal disease is unclear. We performed a Mendelian randomization analysis to examine these associations.

**Methods:** Genetic instruments associated with leisure screen time (LST, an indicator of a sedentary lifestyle) and moderate-to-vigorous intensity physical activity (MVPA) at the genome-wide significance (*P*<5×10^−8^) level were selected from a genome-wide association study (N<703,901). Summary statistics for gastrointestinal diseases were obtained from the UK Biobank study (N>330,000), the FinnGen study (N>220,000), and large consortia. Multivariable MR analyses were conducted for genetically determined LST with adjustment for MVPA and vice versa. We also performed multivariable MR with adjustment for genetically proxied smoking, body mass index (BMI), fasting insulin, and type 2 diabetes for both exposures.

**Results:** Genetically proxied longer LST was associated with increased risk of gastrointestinal reflux, gastric ulcer, duodenal ulcer, chronic gastritis, irritable bowel disease, diverticular disease, Crohn’s disease, non-alcoholic fatty liver disease, alcoholic liver disease, cholecystitis, cholelithiasis, acute pancreatitis, chronic pancreatitis, and acute appendicitis. Most associations remained after adjustment for genetic liability to MVPA. Genetic liability to MVPA was associated with decreased risk of gastroesophageal reflux, gastric ulcer, chronic gastritis, irritable bowel syndrome, cholelithiasis, and acute pancreatitis. The associations attenuated albeit directionally remained after adjusting for genetically predicted LST. Multivariable MR analysis found that BMI and type 2 diabetes mediated the associations of LST and MVPA with several gastrointestinal diseases.

**Conclusion:** The study suggests that a sedentary lifestyle may play a causal role in the development of many gastrointestinal diseases.

## Introduction

Insufficient physical activity affects approximately 27.5% of the global population, with a higher prevalence of 42.3% in high-income countries, significantly contributing to the burden of disease (1). Previous population-based observational studies found that high physical activity levels were associated with a low risk of many gastrointestinal diseases, including gastroesophageal reflux disease (2), esophageal cancer (3), colorectal cancer (4), cholecystitis (5), cholelithiasis (6) and non-alcoholic fatty liver disease (7). The causal potential of the association for liver disease was further supported by a meta-analysis of 11 randomized controlled trials which found that exercise improved hepatic steatosis and serum alanine aminotransferase levels in patients with non-alcoholic fatty liver disease (8). However, the causality of most of these associations is not full established due to potential limitations of observational studies, like residual confounding, misclassification, and reverse causation (9). Sedentary behaviors may also influence human health and the risk caused by a sedentary lifestyle cannot be offset by physical activity (10, 11). Nevertheless, few studies have been conducted to explore the associations between sedentary behavior and the risk of gastrointestinal diseases; and the causal associations of sedentary behavior and physical activity with the risk of developing gastrointestinal diseases remain to be elucidated.

Mendelian randomization (MR) is an epidemiological approach that utilizes genetic variants as instrumental variables to strengthen causal inference (12). Genetic variants are randomly assorted at conception and thus are not subject to confounding. Multivariable MR (MVMR) is an approach of reducing potential pleiotropy or exploring mediation effects by calculating estimates conditionally on the pleiotropic trait or the mediator. Here, we conducted an MR investigation to explore the associations of sedentary behavior and physical activity with a wide range of gastrointestinal diseases.

## Methods

We used leisure screen time (LST) as an indicator of sedentary behavior and moderate-to-vigorous intensity physical activity during leisure time (MVPA) for physical activity. The study design is presented in **Figure 1**. The study was based on data from large genome-wide association studies (GWASs) including the UK Biobank study (13), the FinnGen study (14), the Resource for Genetic Epidemiology Research on Aging (GERA) (15), and the International Inflammatory Bowel Disease Genetics Consortium (IIBDGC) (16). Detailed information on used data sources is shown in **Table S1**. MR estimates were generated separately in each outcome dataset and then combined for each gastrointestinal endpoint using meta-analysis. All included GWASs had been approved by corresponding institutional review boards and ethical committees. All participants had signed informed consent.

**Figure 1.**
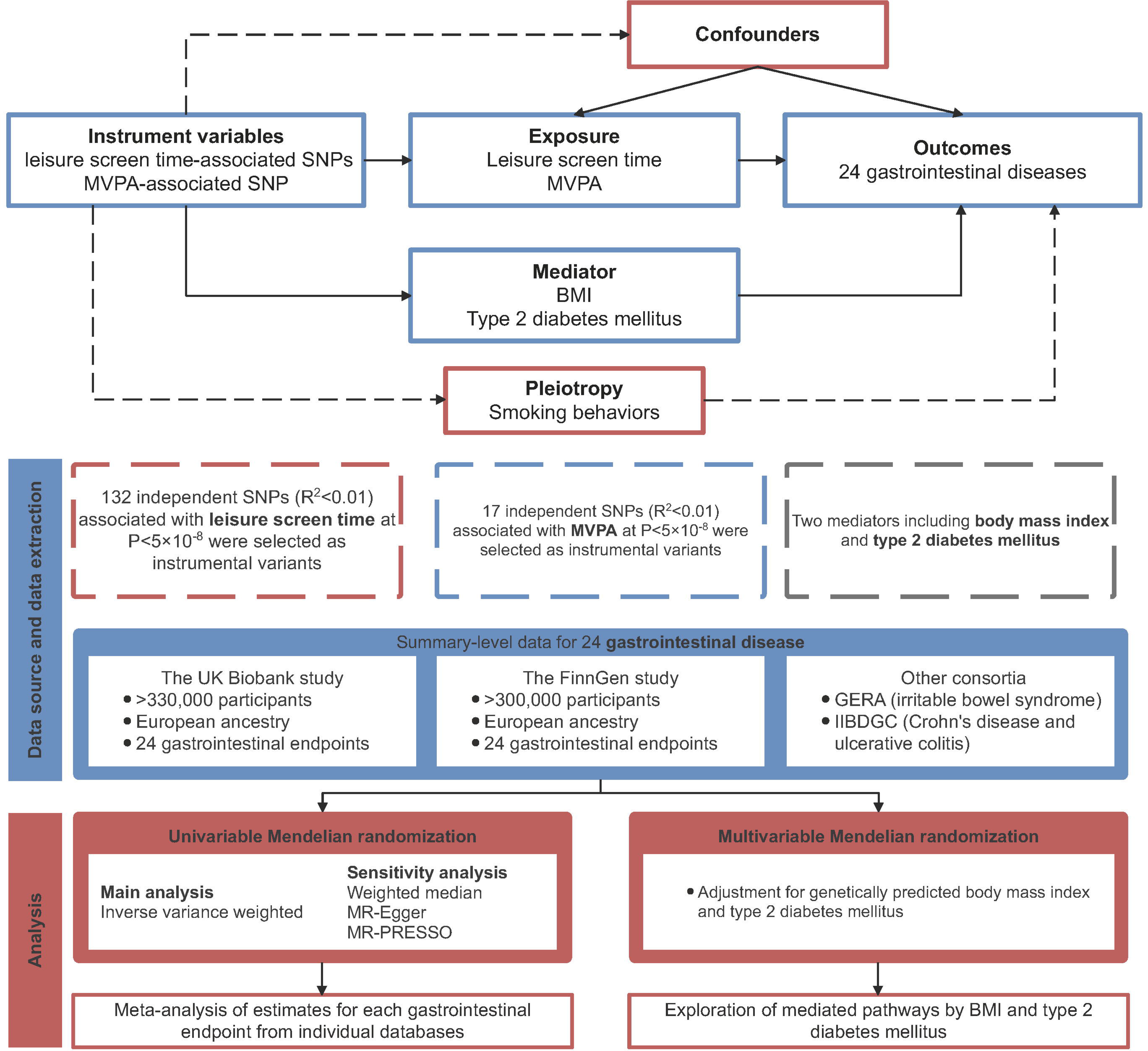
Study design. MVPA, moderate-to-vigorous intensity physical activity during leisure time; MR, Mendelian randomization; GERA Resource for Genetic Epidemiology Research on Aging; IIBDGC, the International Inflammatory Bowel Disease Genetics Consortium; MR-PRESSO, Mendelian randomization pleiotropy residual sum, and outlier.

### Instrumental variable selection

Genetic variants for LST (N=606,820) and MVPA (N=526,725) were extracted from a GWAS meta-analysis of 53 studies of European populations (17). The two phenotypes were based on self-reported data from the questionnaires (17). Single nucleotide polymorphisms (SNPs) at the genome-wide significance threshold (*P* < 5×10^−8^) were identified. These genetic variants were clumped for linkage disequilibrium (defined as r^2^ > 0.01), leaving 132 SNPs as instrumental variables for LST and 17 SNPs for MVPA. Detailed information on used SNPs is shown in **Table S2**.

### Gastrointestinal disease data sources

Genetic associations with 24 gastrointestinal diseases were obtained from the UK Biobank study (18), the FinnGen study (14), and two large consortia, including IIBDGC (16) and GERA (15). The UK Biobank study is a large multicenter cohort study initiated in the United Kingdom between 2006 and 2010 and comprises approximately half a million individuals (18). Summary-level statistics on gastrointestinal outcomes were derived from GWAS summary statistics of European participants from Lee lab (https://www.leelabsg.org/resources) wherein endpoints were defined using codes of the International Classification of Diseases 9th Revision (ICD-9) and ICD-10. Genetic associations were adjusted for sex, birth year, and the first four genetic principal components. We also obtained data on the associations between exposure-SNPs and gastrointestinal diseases from the latest FinnGen study R8 release (14). The FinnGen study combines genetic data from nationwide biobanks and disease status from structured national healthcare databases. The gastrointestinal diseases were defined by the ICD codes (ICD-8, ICD-9, and ICD-10). Genetic associations were adjusted for sex, age, genotyping batch, and 10 principal components. ICD codes used to define the outcomes in UK Biobank and FinnGen are presented in **Table S3**. Additionally, summary-level data from the IIBDGC were extracted for Crohn’s disease (5,956 cases and 14,927 controls) and ulcerative colitis (6,968 cases and 20,464 controls) (16) and summary-level data from the GERA were extracted for irritable bowel syndrome (3,117 cases and 53,520 controls) (15).

### Data sources for smoking, alcohol comsuption, body mass index, diabetic traits

The instrumental variables for smoking initiation, cigarettes smoked per day, alcohol comsuption, body mass index (BMI), fasting insulin and type 2 diabetes mellitus liability were extracted from the corresponding latest publicly available GWASs, respectively (19-22). Summary-level data on smoking initiation from GWAS and Sequencing Consortium of Alcohol and Nicotine use consortium (GSCAN) were extracted where the phenotype reflected smoking status (19). Genetic associations with cigarettes smoked per day were obtained from GSCAN which capture the heaviness of smoking (19). Genetic association with alcohol comsuption were also derive from GSCAN including 941,280 European-ancestry individuals (19). Genetic associations with BMI were obtained from the largest GWAS which containing up to 694 649 individuals of European ancestry (20). Genetic associations with fasting insulin were obtained from a GWAS with 151,013 European-ancestry individuals (21).Summary-level data on type 2 diabetes liability were selected from a meta-analysis GWAS which including 148,726 cases and 965,732 controls (22). Detailed information on these studies is shown in **Table S1**.

### Statistical analysis

The primary MR estimates were calculated using the inverse-variance weighted (IVW) method under multiplicative random-effects. The IVW method provides the most precise estimate under the assumption that all instruments are valid. Estimates for each outcome from different sources were combined using the fixed-effects meta-analysis method. Cochran’s Q value was employed to evaluate the heterogeneity among SNPs’ estimates. Three sensitivity analyses were conducted including the weighted median (23), MR-Egger (24), and Mendelian randomization pleiotropy residual sum and outlier (MR-PRESSO) (25) to examine the robustness of the associations and to detect potential horizontal pleiotropy. The weighted median method provides a consistent estimate if > 50% of the weight comes from valid instrumental variables (23). Although MR-Egger regression provides an estimate usually underpowered, the embedded intercept test can detect potential horizontal pleiotropy (24). MR-PRESSO can identify horizontal pleiotropic outliers and provides a corrected estimate after the removal of the outliers (25).

We performed a sensitivity analysis using genetic variants using a lower linkage disequilibrium threshold of *r*^*2*^=0.001. In addition, given a strong genetic correlation between LST and MVPA (rho=-0.49), we performed multivariable MR (MVMR) analyses with mutual adjustment to explore the independent effects of LST and MVPA on gastrointestinal diseases. To minimize pleiotropy from smoking and alcohol consumption, we performed MVMR with adjustment for genetic predisposition to smoking and alcohol consumption. MVMR were also conducted with adjustment for genetically predicted BMI and type 2 diabetes to explore the mediation of these factors on the associations of LST and MVPA with the risk of gastrointestinal disease.

Power estimation was performed for each gastrointestinal outcome. *F*-statistic was calculated for each SNP and an *F*-statistic > 10 indicated a reliable instrument variable. The conditional *F*-statistic was calculated to characterize instrument strengths in MVMR (Table S5). However, the power analysis and *F*-statistic were not available for MVPA due to its binary phenotype. Odds ratios (ORs) and confidence intervals (CIs) of gastrointestinal diseases were scaled to a one-SD increase in hours/day of genetically predicted LST and a one-unit increase in log odds of genetic liability to MVPA (17). The Benjamini-Hochberg method that controls the false discovery rate (FDR) was applied to correct for multiple testing. The association with a Benjamini–Hochberg adjusted *P* value <0.05 were deemed statistically significant. All analyses were two-sided and performed using the TwoSampleMR (26), MendelianRandomization (23), and MRPRESSO (25) R packages in R software 4.2.2.

## Results

There was no indication of weak instruments for LST (*F*-statistic for each genetic variant was above 10) (**Table S2**). However, not all the conditional F-statistics of instruments were more than 10 in MVMR (**Table S4**). Power calculations showed that there was 80% power to detect the OR ranging from 1.13 to 1.94 for each outcome (**Table S5**).

### LST and gastrointestinal disease

After multiple comparison corrections, genetically predicted longer LST was associated with an increased risk of 14 of 24 gastrointestinal diseases including gastrointestinal reflux, gastric ulcer, duodenal ulcer, chronic gastritis, irritable bowel syndrome, diverticular disease, Crohn’s disease, non-alcoholic fatty liver disease, alcoholic liver disease, cholecystitis, cholelithiasis, acute pancreatitis, chronic pancreatitis, and acute appendicitis (**Figure 2 and Table S6**). The ORs per 1-SD increase in genetically predicted LST ranged from 1.11 (95% CI 1.02, 1.20) for acute appendicitis to 1.62 (95% CI 1.32, 1.99) for non-alcoholic fatty liver disease. Heterogeneity in SNP estimates was detected in the analysis for gastroesophageal reflux, gastric ulcer, chronic gastritis, non-alcoholic fatty liver disease, cholelithiasis, chronic pancreatitis, and acute appendicitis in FinnGen. The associations were directionally consistent in sensitivity analyses (**Table S7**). The MR-Egger intercept test detected potential horizontal pleiotropy in the analysis for gastroesophageal reflux in FinnGen (*P* for intercept <0.05). However, this association persisted in MR-PRESSO analysis after the removal of an outlier (**Table S7**). MR-PRESSO also detected 1-2 outliers in the analyses for gastric ulcer and cholelithiasis; however, these associations remained after removing the outliers (**Table S7**). The associations remained in the sensitivity analysis using a stringent threshold of linkage (**Table S8**). In MVMR adjustment for genetic liability to MVPA, genetically predicted LST was associated with gastroesophageal reflux, gastric ulcer, duodenal ulcer, irritable bowel syndrome, diverticular disease, non-alcoholic fatty liver disease, cholecystitis, cholelithiasis, acute pancreatitis, chronic pancreatitis, and acute appendicitis (*P* < 0.05) although slightly attenuated (**Figure 2** and **Table S9**).

**Figure 2.**
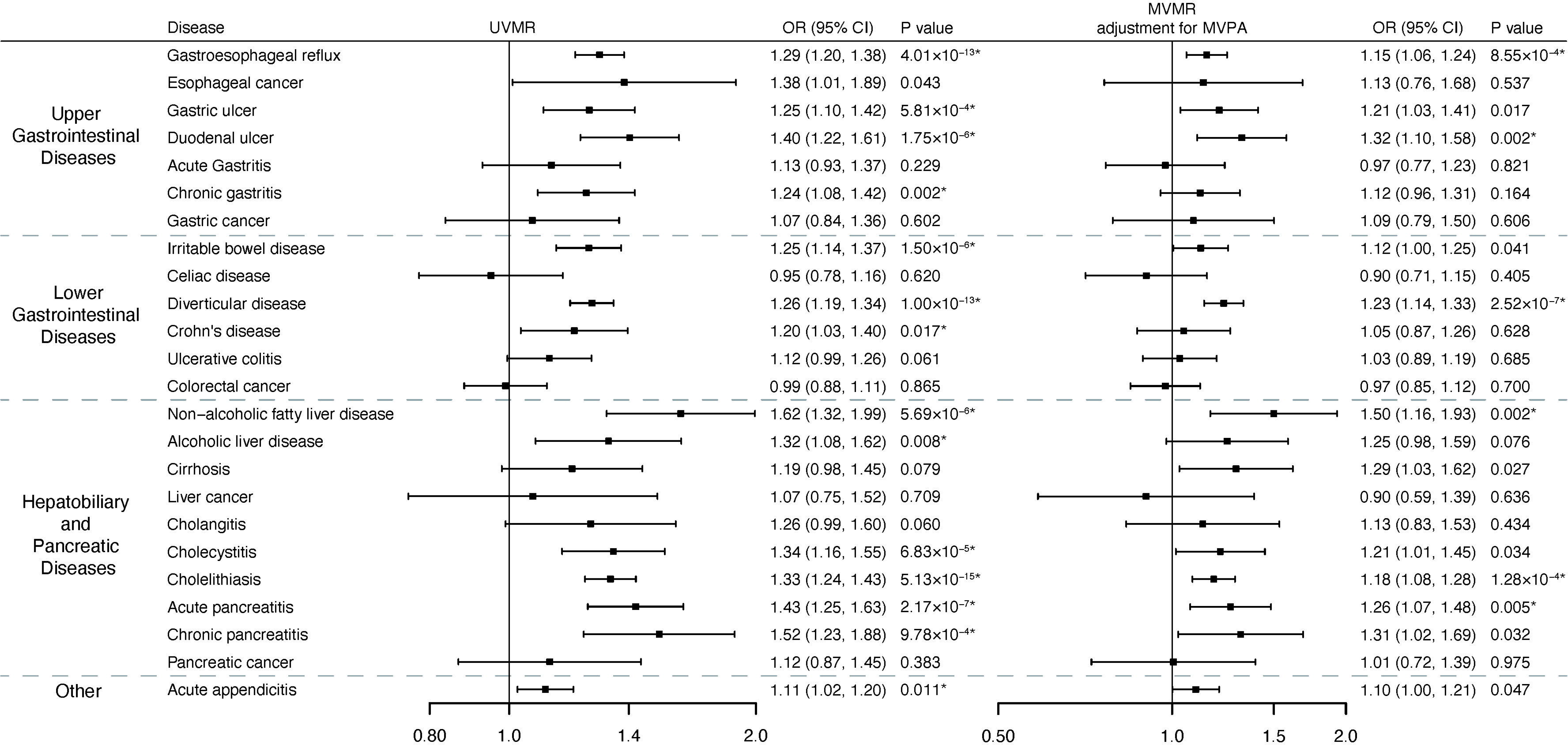
Associations of genetically predicted leisure screen time with 24 gastrointestinal diseases in univariable Mendelian randomization and multivariable Mendelian randomization (adjustment for moderate-to-vigorous intensity physical activity). *Significant association after multiple testing. The estimate of irritable bowel syndrome was meta-analyzed by combining estimates from the UK Biobank study, the FinnGen study, and the Genetic Epidemiology Research on Aging consortium; the estimates of Crohn’s disease and ulcerative colitis were meta-analyzed by combining estimates from the UK Biobank study, the FinnGen study and the International Inflammatory Bowel Disease Genetics Consortium; the estimates of other gastrointestinal disease were meta-analysis by combining estimates from the UK Biobank study and the FinnGen study. ORs for gastrointestinal diseases were scaled to genetically predicted one standard deviation increase in hours/day of leisure screen time.

### MVPA and gastrointestinal diseases

Genetic predisposition to MVPA was associated with a decreased risk of 6 gastrointestinal diseases including gastroesophageal reflux, gastric ulcer, chronic gastritis, irritable bowel syndrome, cholelithiasis, and acute pancreatitis after correcting for multiple testing (**Figure 3 and Table S6**). The ORs of genetically predicted MVPA ranged from 0.49 (95% CI 0.34, 0.71) for acute pancreatitis to 0.72 (95% 0.59, 0.88) for gastroesophageal reflux. There is heterogeneity in SNP estimates for the analysis of gastroesophageal reflux in the UK Biobank (**Table S10**). Estimates across sensitivity analyses were directionally consistent. The MR-Egger intercept test did not detect any indication of pleiotropy. One outlier was identified for gastroesophageal reflux in UK Biobank. The associations persisted after the removal of the outlier (**Table S10**). With a strict linkage disequilibrium threshold, genetically predicted MVPA was still associated with these 6 identified gastrointestinal diseases (**Table S8**). After adjustment for genetically proxied LST, although estimates were not substantially altered, the associations of genetically predicted MVPA with gastric ulcer and chronic gastritis were no longer statistically significant (**Table S9**).

**Figure 3.**
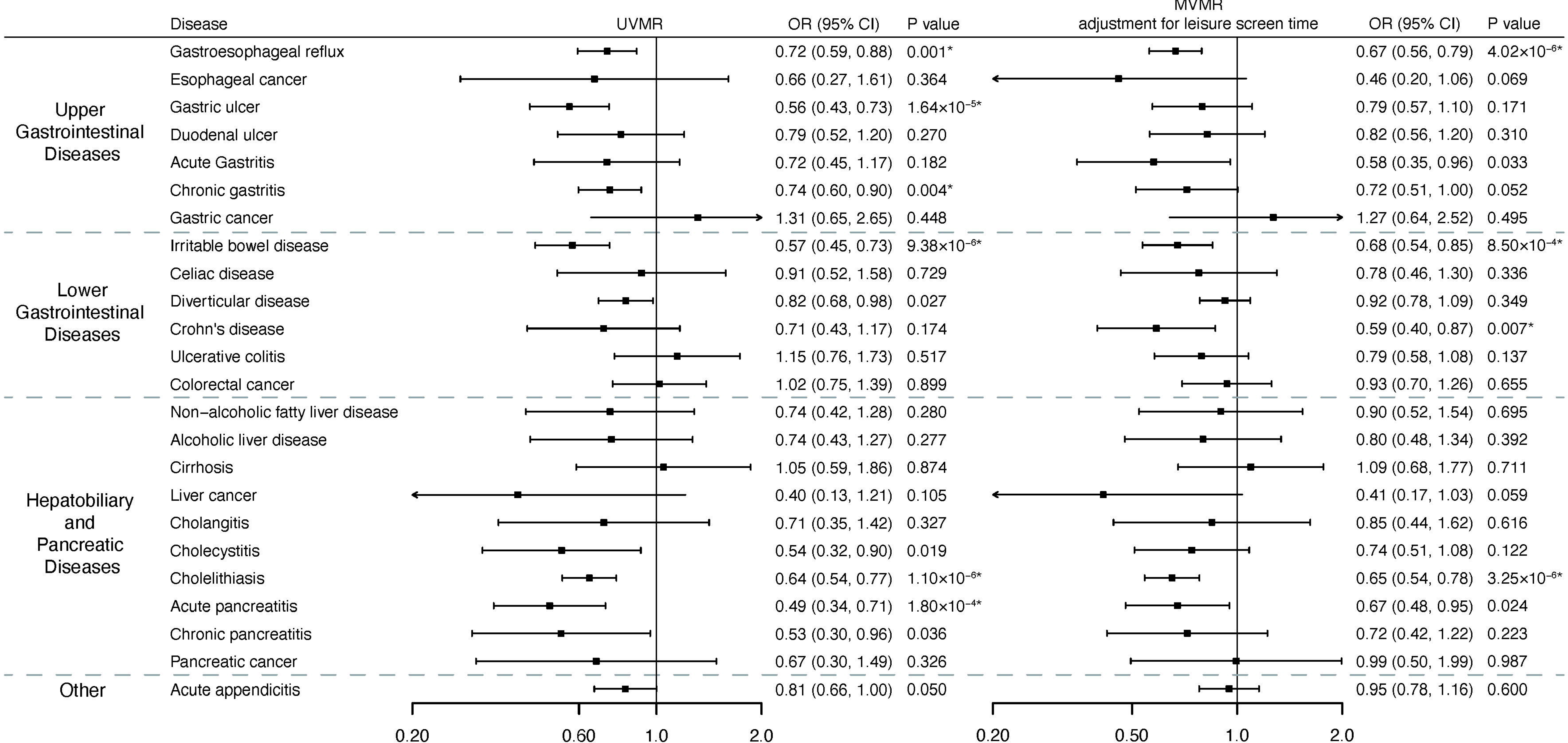
Associations of genetically predicted moderate-to-vigorous intensity physical activity with 24 gastrointestinal diseases in univariable Mendelian randomization and multivariable Mendelian randomization (adjustment for leisure screen time). *Significant association after multiple testing. The estimate of irritable bowel syndrome was meta-analyzed by combining estimates from the UK Biobank study, the FinnGen study, and the Genetic Epidemiology Research on Aging consortium; the estimates of Crohn’s disease and ulcerative colitis were meta-analyzed by combining estimates from the UK Biobank study, the FinnGen study and the International Inflammatory Bowel Disease Genetics Consortium; the estimates of other gastrointestinal disease were meta-analysis by combining estimates from the UK Biobank study and the FinnGen study. ORs for gastrointestinal diseases were scaled to genetically predicted one unit increase in log odds of moderate-to-vigorous intensity physical activity.

### Pleitropic effects of smoking and alcohol consumption

When adjusting for genetically proxied smoking initiation, genetically predicted longer LST was associated with 13 of 24 gastrointestinal diseases except for alcoholic liver disease even though the estimates were modestly attenuated (**Table S11**). Similar results were obtained after adjustment for genetically proxied cigarettes smoked per day (**Table S11**). With adjustment for genetically predicted alcohol consumption, genetically predicted longer LST was still associate with 13 identified gastrointestinal diseases (**Table S11**). After adjusting for genetically proxied smoking initiation, the associations of genetic predisposition to MVPA with gastroesophageal reflux, irritable bowel syndrome, and cholelithiasis remained (**Table S11**). The associations for five gastrointestinal diseases remained significant after adjustment for genetically proxied cigarette smoked per day except for that for chronic gastritis (**Table S11**). Similar findings were observed after adjustment for genetically predicted alcohol consumption (**Table S11**).

### Mediation analysis

The mediation of BMI, fasting insulin, and type 2 diabetes in the associations between LST and 14 identified gastrointestinal diseases is shown in **Table 1 and Table S12**. The proportion of BMI-mediation effect ranged from -1.4% for chronic pancreatitis to 45.6% for cholelithiasis (**Table 1**). The mediation effect of fasting insulin and type 2 diabetes was less pronounced than that of BMI. The proportion of the mediation effect of fasting insulin ranged from -1.3% for Crohn’s disease to 15.7% for non-alcoholic fatty liver disease, and the proportion of mediation effect of type 2 diabetes ranged from -1.2% for Crohn’s disease to 18.4% for non-alcoholic fatty liver disease (**Table 1**).

**Table 1.** Proportion of mediation effect in association between leisure screen time and 14 gastroinestinal disease via each mediator

| Disease/Mediator | Body mass index | Fasting insulin | Type 2 diabetes |
| --- | --- | --- | --- |
| Gastroesophageal reflux | 13.33% | 4.23% | 4.09% |
| Gastric ulcer | 10.80% | 9.05% | 8.42% |
| Duodenal ulcer | 13.70% | 7.85% | 9.27% |
| Chronic gastritis | 3.99% | 11.98% | 13.18% |
| Irritable bowel disease | -2.17% | 4.28% | 4.66% |
| Diverticular disease | 14.73% | 3.29% | 4.18% |
| Crohn's disease | 11.19% | -1.28% | -1.20% |
| Non-alcoholic fatty liver disease | 32.52% | 15.68% | 18.41% |
| Alcoholic liver disease | 11.20% | 6.63% | 5.38% |
| Cholecystitis | 37.52% | 8.14% | 7.66% |
| Cholelithiasis | 45.63% | 8.92% | 9.01% |
| Acute pancreatitis | 12.78% | 3.42% | 3.65% |
| Chronic pancreatitis | -1.43% | 4.64% | 5.36% |
| Acute appendicitis | 27.41% | 2.13% | 1.74% |

The results of the mediation analysis of BMI, fasting insulin, and type 2 diabetes in association between MVPA and gastrointestinal diseases are shown in **Table 2 and Table S12**. BMI mediated around 39.1% of the effect of MVPA on cholelithiasis (**Table 2**). Fasting insulin mediated 2.7%-10.3% effect of MVPA on 6 gastrointestinal diseases. The mediation effect of type 2 diabetes ranged from 3.78% for gastroesophageal reflux to 18.1% for chronic gastritis (**Table 2**).

**Table 2.** Proportion of mediation effect in association between moderate-to-vigorous intensity physical activity and 15 gastroinestinal disease via each mediator

| Disease/Mediator | Body mass index | Fasting insulin | Type 2 diabetes |
| --- | --- | --- | --- |
| Gastroesophageal reflux | 7.40% | 4.26% | 3.78% |
| Gastric ulcer | 3.59% | 5.78% | 5.79% |
| Chronic gastritis | 2.79% | 10.33% | 18.11% |
| Irritable bowel disease | 1.80% | 2.72% | 3.90% |
| Cholelithias | 39.13% | 6.84% | 10.89% |
| Acute pancreatitis | 11.11% | 3.91% | 6.44% |

## Discussion

This study comprehensively investigated the causal associations of LST and MVPA with the risk of 24 gastrointestinal diseases using MR analysis. Genetically proxied longer LST was associated with an increased risk of 14 gastrointestinal diseases. Genetic liability to MVPA was inversely associated with 6 gastrointestinal diseases. The associations for LST remained overall stable after adjustment for genetic liability to MVPA and vice versa. However, some associations attenuated after adjustment for genetically proxied smoking behaviors, which indicates the pleiotropic effects of smoking in this analysis. Multivariable MR analysis also found that genetically proxied BMI, fasting insulin levels, and type 2 diabetes partially mediated the associations of LST and MVPA with several gastrointestinal diseases.

The current MR found evidence that genetically predicted LST and MVPA were associated with several upper and lower gastrointestinal diseases. Previous observational studies suggested that high levels of physical activity were associated with a low risk of gastroesophageal disease (2, 27), gastric ulcer (28), duodenal ulcer (29), diverticular disease (30, 31) and that lack of habitual physical exercise was associated with a higher risk of gastroesophageal reflux (27). Our MR study supported these findings and further corroborated the causality of these associations.

We also provided evidence that genetically predicted LST was positively associated with some hepatobiliary and pancreatic outcomes including non−alcoholic fatty liver disease, alcoholic liver disease, cholecystitis, cholelithiasis, pancreatitis, and acute appendicitis. Genetically predicted MVPA was inversely associated with cholecystitis, cholelithiasis, and acute pancreatitis. A prospective cohort study including 360,047 participants showed that longer sedentary time was associated with a higher risk of chronic liver disease and replacing 1 h/day sedentary time with equivalent physical activity was associated with a lower risk of chronic liver disease (30), which was in line with our findings. Based on findings from the Nurses’ Health Study, sedentary behavior was independently associated with an increased risk of cholecystectomy (32). Extending the previous observational findings, our MR investigation strengthened the evidence that physical activity was associated with a low risk of cholecystitis (5) and cholelithiasis (6). Our MR investigation also confirmed results from previous MR studies where genetically predicted longer television watching time was associated with higher levels of hepatic fat (33). Another MR study demonstrated that genetically predicted vigorous physical activity was associated with a decreased risk of non-alcoholic fatty liver disease (34). Our findings also provide novel evidence on the associations of LST and MVPA with acute appendicitis which need to be verified.

We did not observe positive associations of genetically predicted LST or MVPA with colorectal cancer or esophageal cancer, which was identified in previous observational studies (35, 36). Besides, a previous MR analysis comprising 31,197 colorectal cases found that genetically predicted MVPA was associated with a decreased risk of colorectal cancer (37). This discrepancy is likely caused by a relatively small sample size of these cancers in our MR analysis.

Our results also provided information that genetically predicted longer LST was positively associated with the risk of several gastrointestinal diseases independent of MVPA and vice versa. These findings suggest that reducing sedentary time and increasing physical activity were both important for gastrointestinal disease prevention. Besides, after mutually adjusting for LST and MVPA, the association for chronic gastritis became null which could be explained by the similar constructs picked by LST and MVPA. Thus, the insignificant associations after mutual adjustment did not necessarily mean that either LST or MVPA was not a risk factor for chronic gastritis. In MVMR adjusting for smoking initiation and cigarettes smoked per day, some identified associations attenuated which indicated the potential horizontal pleiotropy introduced by smoking behaviors. Of note, we observed that BMI, fasting insulin, and type 2 diabetes seemed to partly mediate the associations of LST and MVPA with gastrointestinal diseases, which indicates that metabolic factors may involve in the pathological process. In addition, given that a comparative proportion of the effect was mediated by type 2 diabetes, our results support the current guidelines that recommend increasing activity and decreasing sedentary behavior among type 2 diabetes patients from the American Diabetes Association to improve their overall well-being (38). Similarly, more physical exercise and less sedentary time should also be recommended among obese individuals as a part of the management of gastrointestinal health due to the mediation effects of BMI observed in the link from physical activity and sedentary lifestyle to gastrointestinal disease in this study.

Mechanisms linking sedentary behavior and physical activity with the gastrointestinal disease are not completely understood. Apart from being overweight, animal experiments have suggested that prolonged sitting might disturb lipid metabolism and result in the reduction of lipoprotein lipase activity which then raises the possibility of impaired metabolic actions (39, 40). Besides, there was evidence that sedentary behavior might lead to increased systemic chronic inflammation, which may also increase the risk of developing gastrointestinal diseases (41). In addition, high levels of physical activity and a shorter sedentary time have been associated with lower insulin levels (42), which have been indicated to play a role in the development of many gastrointestinal diseases (43-45). The diversity, composition, and functionality of gut microbiota can be modified by physical exercise possibly via intestinal barrier preservation and bile acid homeostasis improvement (46). In addition, standing up benefits physiological processes including postural blood flow, energy expenditure, and muscle contraction, which may promote glucose regulation, mitochondrial function, and endothelial function (47). This may explain why adjustment for MVPA barely affected the positive association between LST and several gastrointestinal diseases.

Compared to physical activity, sedentary behaviors in relation to gastrointestinal diseases were less been investigated. Our findings on LST are potentially relevant to interventions and guidance aimed at reducing gastrointestinal diseases through behavioral intervention. Besides, LST has increased by approximately 1 hour for young children and 0.7 hours for adults during the COVID-19 pandemic (48), which may exacerbate these harms. According to our findings, emphasizing and adopting multilevel measures are critical to preventing unhealthy screen time.

The major strength of the present study is the MR design, which minimized bias from confounding and reverse causality and thus improved the causal inference in the associations of LST and MVPA with gastrointestinal diseases. We also used several independent outcome sources and combined the estimates, which increased statistical power as well as strengthened our findings by the observed consistency of results. Another strength is that we confined our analysis to individuals of European ancestry, which minimized the population stratification bias.This study also has several limitations.

A major limitation of the current MR study was horizontal pleiotropy, which could affect the validity of our MR findings. However, we conducted a series of sensitivity analyses including adjustment for two smoking behaviors, and the consistent results indicated limited pleiotropy. Another limitation is the relatively limited sample size in several gastrointestinal diseases, which means this study might have had inadequate power to detect weak associations between the exposure and diseases. A relatively small number of instruments is available for MVPA which may result in inadequate statistical power and underestimating the impact of MVPA on gastrointestinal diseases. Besides, our study participants were of European ancestry, and thus the observed associations may not be directly generalizable to other populations. A further potential limitation is that individuals of the UK Biobank were included in both the exposure and outcome datasets, which might introduce some bias in the causal estimates in the direction of the estimates of observational studies. However, we employed non-overlapping datasets (e.g., the FinnGen study) and the consistent results indicated that the bias due to sample overlap is reasonably small. The conditional F-statistics in MVMR indicated possible weak instrument bias. However, the small sample overlap minimized the possibility of this bias. Finally, the LST and MVPA were self-reported which could be biased by recall and awareness of the beneficial effects of physical activity.

In conclusion, this MR investigation found that genetically proxied longer LST was associated with an increased risk of 14 gastrointestinal diseases, whereas genetic liability to MVPA is associated with a decreased risk of 6 gastrointestinal diseases. Considering that physical activity and sedentary behavior are inversely correlated, our findings support the independent benefits of reducing sedentary time and promoting physical acidity in gastrointestinal disease prevention.Future studies are encouraged to explore the joint effect of LST and MVPA on gastrointestinal health.

## Data Availability

Data can be obtained upon a reasonable request to corresponding authors.

## Statements

### Contributorship

All authors read and approved the final manuscript and author contributions statement using CRediT with the degree of contribution:

Jie Chen (Conceptualization: Equal; Methodology: Equal; Formal analysis: Equal; Data curation: Equal; and Writing - review & editing: Equal)

Xixian Ruan (Conceptualization: Equal; Methodology: Equal; Formal analysis: Equal; Data curation: Equal; and Writing – original draft: Equal)

Tian Fu (Conceptualization: Equal; Methodology: Equal; Formal analysis: Equal; Data curation: Equal; and Writing – original draft: Equal)

Shiyuan Lu (Conceptualization: Supporting; Writing - review & editing: Supporting) Dipender Gill (Conceptualization: Supporting; Methodology: Supporting; Writing - review & editing: Equal)

Zixuan He (Conceptualization: Supporting; Writing - review & editing: Supporting)

Stephen Burgess (Conceptualization: Supporting; Methodology: Supporting; Writing - review & editing: Equal)

Edward L Giovannucci (Conceptualization: Supporting; Methodology: Supporting; Writing - review & editing: Equal)

Susanna C. Larsson (Conceptualization: Equal; Methodology: Equal; Data curation: Equal; and Writing - review & editing: Leading)

Minzi Deng (Conceptualization: Equal; Data curation: Equal; and Funding acquisition: Equal; Writing - review & editing: Equal)

Shuai Yuan (Conceptualization: Leading; Data curation: Equal; Writing - review & editing: Leading)

Xue Li (Conceptualization: Equal; Data curation: Equal; Funding acquisition: Leading; and Writing - review & editing: Leading)

### Data Sharing

Data can be obtained upon a reasonable request to corresponding authors.

