## Supplementary material for "Sedentary lifestyle, Physical Activity, and Gastrointestinal Diseases: Evidence from Mendelian Randomization Analysis": Table S1-TableS12

Table S1. Information of included studies and consortia.

Table S2. Single nucleotide polymorphisms used as instrumental variables for leisure screen time and moderate-to-vigorous intensity physical activity during leisure time

Table S3. Number of cases and definition of gastrointestinal diseases in UK Biobank and FinnGen

Table S4. Conditional F statistics from multivariable MR

Table S5 Power estimation of current Mendelian randomization analysis

Table S6. False discovery rate adjusted p values for all tested associations in main analysis

Table S7. Estimates of genetical liability to leisure screen time on gastrointestinal disease in univariable mendelian randomization

Table S8. Association of leisure screen time and moderate-to-vigorous intensity physical activity during leisure time with gastrointestinal disease in univariable mendelian randomization using inverse variance weighted method (linkage disequilibrium threshold of r2=0.001)

Table S9. Association of genetically predicted leisure screen time and moderate-to-vigorous intensity physical activity during leisure time on gastrointestinal diseases in multivariable Mendelian randomization

Table S10.Estimates of genetical liability to moderate-to-vigorous intensity physical activity during leisure time on gastrointestinal disease in univariable mendelian randomization

Table S11.Estimates of genetical liability to leisure screen time and moderate-to-vigorous intensity physical activation gastrointestinal disease adjusting for smoking phenotype

Table S12. Estimates of leisure screen time and moderate-to-vigorous intensity physical activity on gastrointestinal diseases mediated by four multiple mediators

Figure S1. Summary of associations of genetically predicted leisure screen time and moderate-to-vigorous physical activity with 24 gastrointestinal diseases.

All tables and figure can be obtained in OSF data respiratory:

<https://osf.io/xr45h/?view_only=35edb8b253cc45d7a444bc15bc141a7c>

Please view the file by clicking the xlsx file.

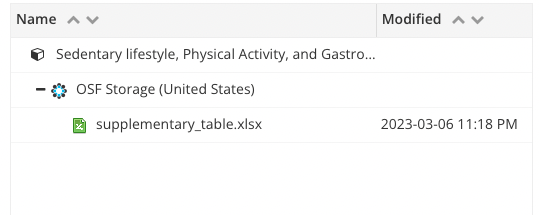
